# Maternal health in rural Pakistan: An analysis of knowledge and practices of antenatal care using the socio-ecological model

**DOI:** 10.1101/2024.03.31.24305141

**Authors:** Nadia Agha, Rahim Dad Rind, Shoukat Ali Mahar, Moomal Mendhri Channa, Fizza Ansar, Sadiq Bhanbhro

## Abstract

**Background:** Pakistan carries a heavy burden of maternal and child health vulnerabilities. Maternal health can be prevented with culturally specific interventions and women’s enhanced access to healthcare facilities, yet women in rural areas in Pakistan are deprived of such interventions. In this study, we assessed the prevalent knowledge and practices of antenatal care and the impact of low-income and gender inequalities on maternal health in rural areas of Pakistan.

**Method:** We conducted a cross-sectional study using the Socio-Ecological Model (SEM) in five villages in the Sindh province of Pakistan. We tested hypotheses using descriptive reliability and regression techniques.

**Results:** Findings show that the socio-geographical and cultural factors influencing maternal health were embedded within the different levels of SEM. The prevalence of Maternal Health Vulnerabilities (MHV) was high in villages where women had limited literacy and awareness. Of the 352 participants, 173 (49%) women were not literate and could not read pregnancy-related information imparted through leaflets or brochures. Data showed that women consumed less milk, multivitamins, and proper food due to poverty. At the same time, they had restricted access to health facilities: 119 (34%) women had lost their babies after birth. In this study, poverty and gender inequality correlate with MHV (significant 65.60% and .000 and 10.50% and .000, respectively) and exacerbate the situation by complementing each other.

**Conclusion:** Culturally appropriate and evidence-based interventions are needed to reduce gender inequality and prioritise women’s and girls’ maternal health needs. This study has considered cultural, social, geographical, and economic factors influencing maternal health. It provides a nuanced account of a complex web of barriers to attaining Sustainable Development Goals (SDGs), particularly goals 3, ‘Good health and well-being’ and 5 ‘, Gender equality’.

## Introduction

### Background

Maternal and child mortality is a crucial health challenge prevailing in many low and middle-income countries (LMICs) [1]. Worldwide, around 800 women died every day in 2020 from pregnancy and childbirth-related complications [2], making the global Maternal Mortality Rate (MMR) 216 deaths per 100,000 births [3]. It is estimated that 94% of maternal mortality occurs in LMICs [4].

Globally, South Asia and Sub-Saharan African countries carry the burden of maternal deaths [5]. Maternal mortality in these regions reached up to 87% of the total global burden [2]. About 80% of these cases could have been prevented if women were given access to basic healthcare facilities [6]. Pakistan is one of the leading South Asian countries in terms of high MMR [7]. It carries a more significant burden of maternal, foetal and child mortality rates [8]. The MMR in Pakistan is 186 deaths per 100,000 live births, which is 26% higher in rural than in urban areas [9].

There are myriad factors contributing to maternal and perinatal health vulnerabilities. The leading factors of pregnancy-related mortality and morbidity are low household income, low education, poor healthcare services, gender-based disparities, socio-cultural values, more considerable distance, malnutrition, unfair resource distribution and political environment. These are significant indicators for rising maternal mortality cases in rural areas [7]. Unmet nutritional needs, heavy physical labour, infections, and some other primary chronic conditions like being overweight and diabetes further deteriorate pregnancy outcomes [10].

Maternal mortality can be prevented with access to increased professionally delivered interventions, more specifically, coverage of crucial obstetric care, access to safe abortion services, active management during the third stage of labour and the application of anticonvulsants for women with pre-eclampsia [11]. The knowledge and implementation of Antenatal Care (ANC) can save the lives of many women. For this, routine antenatal nutrition, maternal and foetal checkups, preventive measures, intervention for managing common physiologic symptoms in pregnancy, and health-related interventions significantly improve the quality of ANC and its utilisation [12].

Pakistan is a signatory of the Sustainable Development Goals (SDGs) Agenda 2030, but it still lags as the MMR is higher in rural settings. The quality of healthcare is a crucial concern for the coverage of universal health and equity and for reducing the occurrence of maternal and newborn mortalities [13]. National programmes and interventions often fail due to distance and poor infrastructure, more specifically for the people living in rural areas [14]. Therefore, it is essential to understand the existing ANC practices that lead to Maternal Health Vulnerabilities (MHV), particularly in less privileged areas. This study was designed to understand how poverty and gender inequality influence maternal health outcomes in rural areas of Pakistan and to suggest the areas where interventions are needed. This is a quantitative study on women of reproductive age that assesses the existing ANC-related practices, women’s access to healthcare services, and the barriers they experience in accessing these services in rural areas in Sindh, Pakistan.

We aimed to identify socio-cultural and geographical barriers to maternal health in rural areas of Sindh. Research in these areas has not explored and analysed these factors. The study sought to evaluate the prevalent knowledge and practices of antenatal care and the impact of low-income and gender inequality on maternal health in rural areas of Pakistan.

## Methods

This is a cross-sectional study exploring MHV in underserved areas of Sindh. The study was conducted between November 2023 and mid-February 2024 in five villages of Taluka Sukkur. A random sampling technique was used to select households in the villages. We recorded the recent and ongoing ANC practices followed by rural women and how their household income and gender inequality influenced those practices.

### Theoretical framework

The Socio-Ecological Model (SEM) has been applied to this study to assess MHV in rural areas of Sindh, Pakistan. This model has been used extensively to understand individuals’ behaviours about health [15, 16, 17]. According to SEM, an individual has a reciprocal relationship with their environment. The model acknowledges social, cultural, and environmental factors and their associations with biological factors [18]. The model used in this study was borrowed from the [19, 20] ecological model, which consists of four levels influencing the conditions and behaviours about health. The SEM suggests four levels, i.e., microsystem, mesosystem, exosystem and macrosystem. It considers the interplay between individuals and their settings and recognises the dynamic network of factors that shape an individual’s behaviour on the individual, community, and societal levels [21]. SEM is applied to this study to record and analyse the different barriers women experience at these four levels in accessing ANC services.

**Fig 1.**
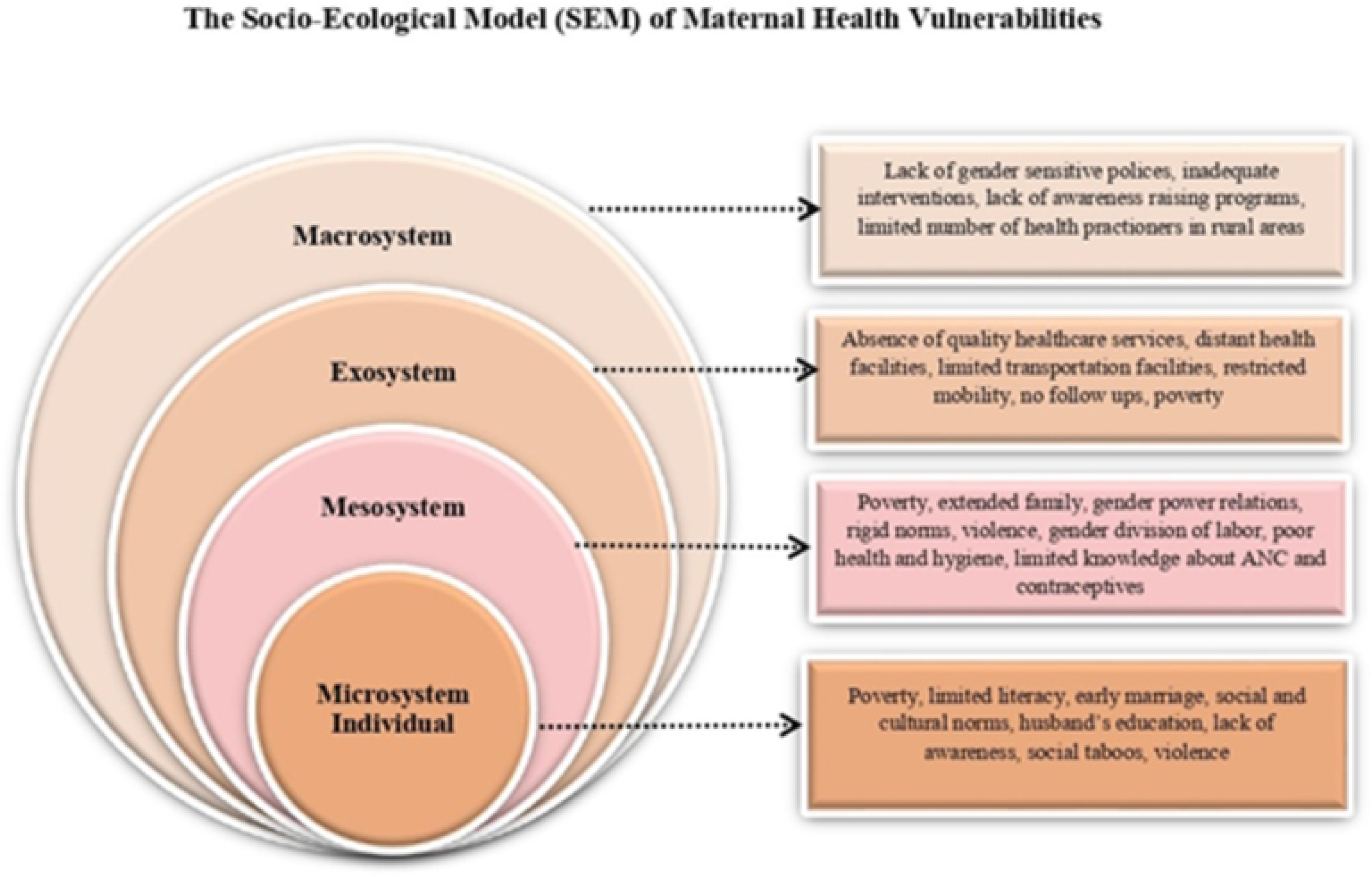
Application of (SEM) Socio-Ecological Model to assess maternal health vulnerabilities.

### Data collection

For this study, we surveyed 360 households and collected data from 352 women of reproductive age. Participants who met the inclusion criteria were selected, i.e., a) women of reproductive age, b) delivered babies within the last two years, and c) residents of rural and less privileged areas. A well-structured, closed-ended questionnaire was prepared after reviewing the literature on the subject. The questionnaire included various themes, including demographic characteristics, child-related variables, and knowledge and practices of ANC-related questions. The first part included demographic variables such as age, marital status, husband and wife’s education, occupation, monthly income, and total number of children. The child-related factors were also included in the first part, such as the child who died after birth (yes/no), the number of children who died after birth, the cause of child mortality, and the age of the children at death.

The second part included a 24-item scale which included questions related to knowledge and practices of ANC among women.

### Ethical approval

The Research Ethics Committees of Aga Khan University, Karachi, and Sheffield Hallam University approved this study. We followed all ethical protocols mentioned in the proposal during the data collection. For example, prior informed and verbal consent was sought from the participants as most of the women (49%) were not literate at all, and 19% had only primary-level education. All the participants were briefed about the study before collecting the data. They were assured of keeping their identity confidential and anonymising all identifiable information.

### Statistical analysis

Data was analysed using SPSS version 24. This study carried out three variables, comprised of 24 questions in which demographic, descriptive reliability and regression techniques were used to test the hypotheses.

## Results

### Descriptive Analysis

Table 1 summarises the demographic characteristics of the study participants. Out of the 352 women of reproductive age, most (21%) belonged to the age group of 18-22, 22% were between 23-27, 24% were between 28-32, and 18% belonged to the age group of 33-37. The marital status variable was taken as married, divorced, or widowed.

**Table 1.**
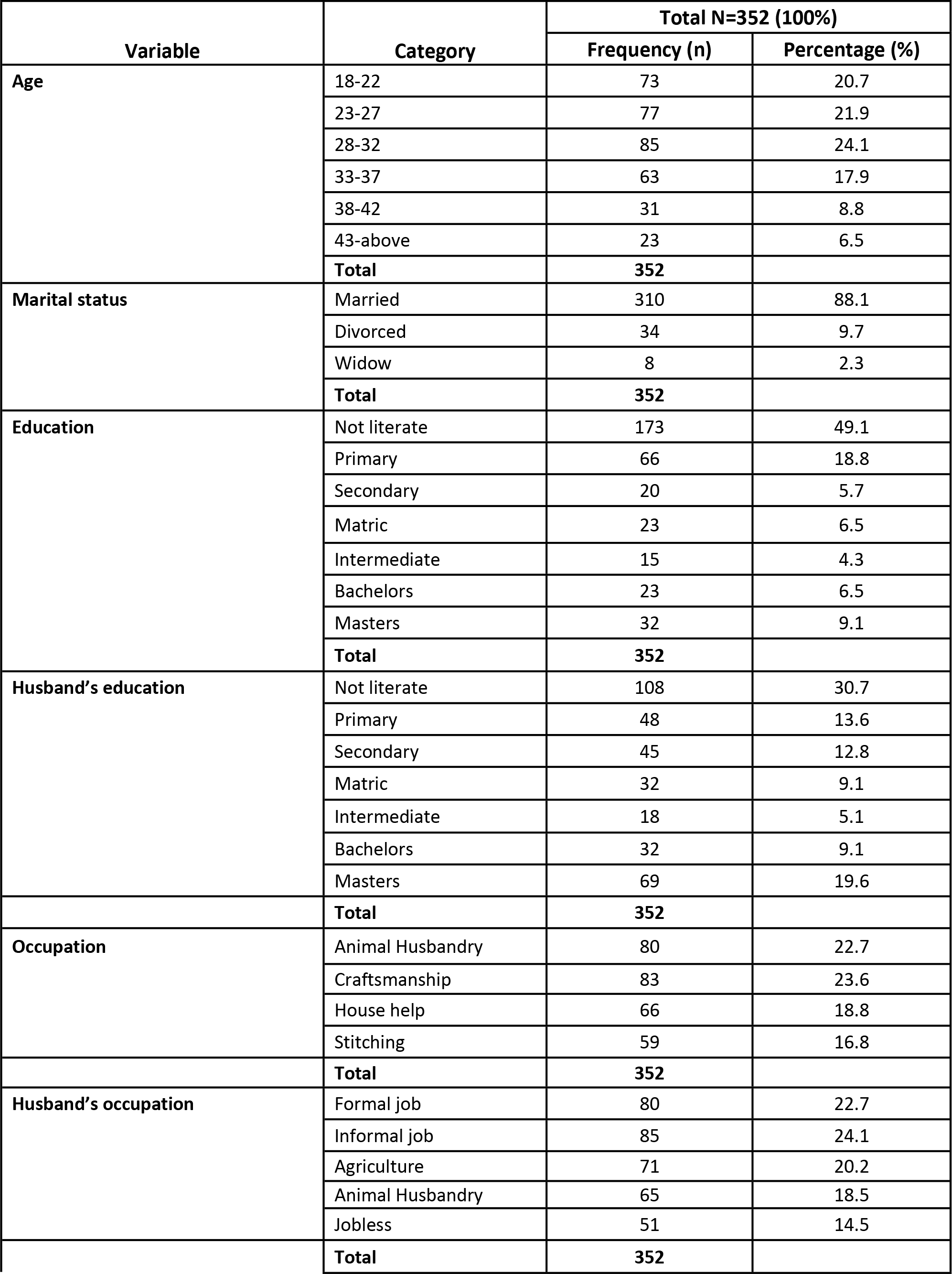
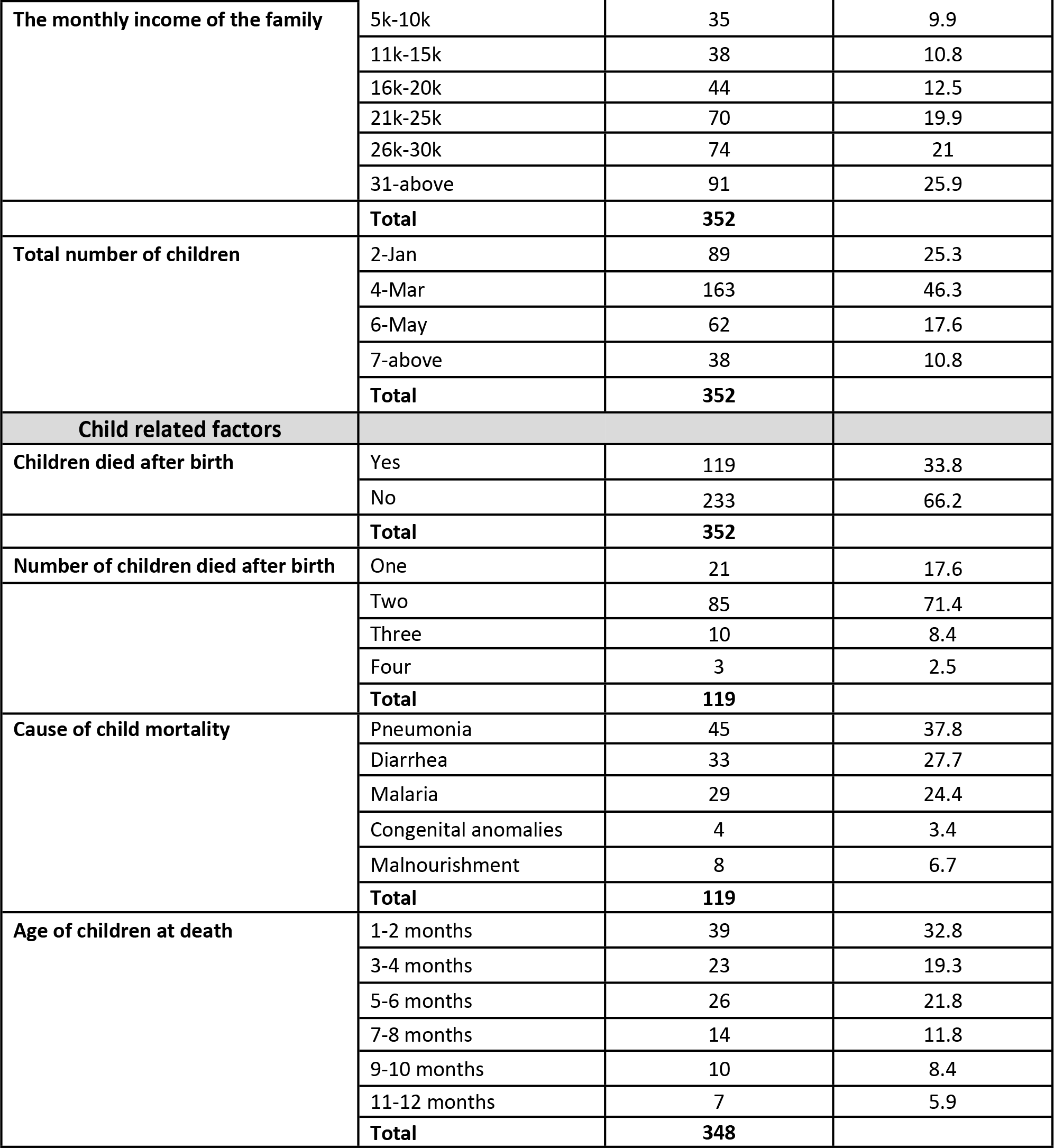
Demographics of participants and child-related factors.

| Variable | Category | Total N=352 (100%) |  |
| --- | --- | --- | --- |
|  |  | Frequency (n) | Percentage (%) |
| Age | 18-22 | 73 | 20.7 |
|  | 23-27 | 77 | 21.9 |
|  | 28-32 | 85 | 24.1 |
|  | 33-37 | 63 | 17.9 |
|  | 38-42 | 31 | 8.8 |
|  | 43-above | 23 | 6.5 |
|  | **Total** | **352** |  |
| Marital status | Married | 310 | 88.1 |
|  | Divorced | 34 | 9.7 |
|  | Widow | 8 | 2.3 |
|  | **Total** | **352** |  |
| Education | Not literate | 173 | 49.1 |
|  | Primary | 66 | 18.8 |
|  | Secondary | 20 | 5.7 |
|  | Matric | 23 | 6.5 |
|  | Intermediate | 15 | 4.3 |
|  | Bachelors | 23 | 6.5 |
|  | Masters | 32 | 9.1 |
|  | **Total** | **352** |  |
| Husband's education | Not literate | 108 | 30.7 |
|  | Primary | 48 | 13.6 |
|  | Secondary | 45 | 12.8 |
|  | Matric | 32 | 9.1 |
|  | Intermediate | 18 | 5.1 |
|  | Bachelors | 32 | 9.1 |
|  | Masters | 69 | 19.6 |
|  | **Total** | **352** |  |
| Occupation | Animal Husbandry | 80 | 22.7 |
|  | Craftsmanship | 83 | 23.6 |
|  | House help | 66 | 18.8 |
|  | Stitching | 59 | 16.8 |
|  | **Total** | **352** |  |
| Husband's occupation | Formal job | 80 | 22.7 |
|  | Informal job | 85 | 24.1 |
|  | Agriculture | 71 | 20.2 |
|  | Animal Husbandry | 65 | 18.5 |
|  | Jobless | 51 | 14.5 |
|  | **Total** | **352** |  |
| **The monthly income of the family** | 5k-10k | 35 | 9.9 |
|  | 11k-15k | 38 | 10.8 |
|  | 16k-20k | 44 | 12.5 |
|  | 21k-25k | 70 | 19.9 |
|  | 26k-30k | 74 | 21 |
|  | 31-above | 91 | 25.9 |
|  | **Total** | **352** |  |
| **Total number of children** | 2-Jan | 89 | 25.3 |
|  | 4-Mar | 163 | 46.3 |
|  | 6-May | 62 | 17.6 |
|  | 7-above | 38 | 10.8 |
|  | **Total** | **352** |  |
| **Child related factors** |  |  |  |
| **Children died after birth** | Yes | 119 | 33.8 |
|  | No | 233 | 66.2 |
|  | **Total** | **352** |  |
| **Number of children died after birth** | One | 21 | 17.6 |
|  | Two | 85 | 71.4 |
|  | Three | 10 | 8.4 |
|  | Four | 3 | 2.5 |
|  | **Total** | **119** |  |
| **Cause of child mortality** | Pneumonia | 45 | 37.8 |
|  | Diarrhea | 33 | 27.7 |
|  | Malaria | 29 | 24.4 |
|  | Congenital anomalies | 4 | 3.4 |
|  | Malnourishment | 8 | 6.7 |
|  | **Total** | **119** |  |
| **Age of children at death** | 1-2 months | 39 | 32.8 |
|  | 3-4 months | 23 | 19.3 |
|  | 5-6 months | 26 | 21.8 |
|  | 7-8 months | 14 | 11.8 |
|  | 9-10 months | 10 | 8.4 |
|  | 11-12 months | 7 | 5.9 |
|  | **Total** | **348** |  |

### Reliability

The reliability of the knowledge and practices questionnaire was assessed by using Cronbach’s alpha. Table 2 represents the cumulative alpha value as .844, suggesting a relatively acceptable internal consistency as the Cronbach alpha results at 0.7 and 0.8 are considered excellent. All the items had corrected item-total correlations. However, .95 or greater values are creating a disturbance in internal consistency [22]. The Cronbach alpha value exists between 0-1.

**Table 2.** Corrected item-total correlation and Cronbach’s alpha values of the 24 items of Knowledge and Practices of ANC.

| Item-Total Statistics | Scale<br>Mean if<br>Item<br>Deleted | Scale<br>Variance if<br>Item<br>Deleted | Corrected<br>Item-Total<br>Correlation | Cronbach's<br>Alpha if<br>Item<br>Deleted |
| --- | --- | --- | --- | --- |
| I have less consumption of milk during my last pregnancy. | 123.8182 | 329.112 | 0.225 | 0.843 |
| I did not take a proper diet during my last pregnancy. | 124.4176 | 325.281 | 0.315 | 0.841 |
| I have to ask for money needed for my treatment from my husband. | 124.1506 | 329.325 | 0.203 | 0.844 |
| I am exposed to violence due to my husband's unemployment during pregnancy. | 124.3324 | 327.288 | 0.253 | 0.842 |
| My household responsibilities do not change during pregnancy. | 124.1903 | 330.616 | 0.156 | 0.845 |
| Household responsibilities quickly made me tired during pregnancy. | 124.2102 | 307.158 | 0.573 | 0.833 |
| I prefer homebirths due to the lack of money. | 124.3097 | 310.14 | 0.495 | 0.835 |
| Healthcare facilities are affordable in government hospitals. | 123.8324 | 315.815 | 0.432 | 0.837 |
| Access to standard healthcare services is difficult. | 124.2301 | 303.511 | 0.606 | 0.831 |
| The source of water and access to sanitation is poor. | 124.0256 | 307.119 | 0.586 | 0.832 |
| I get permission to visit the hospital from my husband during pregnancy. | 124.2216 | 308.264 | 0.531 | 0.834 |
| I do not know much about modern healthcare facilities. | 124.25 | 305.556 | 0.552 | 0.833 |
| My husband decides about family planning. | 124.0653 | 307.457 | 0.559 | 0.833 |
| I do not seek healthcare facilities during pregnancy if there is no companion. | 124.2585 | 309.098 | 0.508 | 0.835 |
| I do not make the decision as to which hospital to go to for a checkup during pregnancy. | 123.6932 | 331 | 0.197 | 0.844 |
| I cannot visit the health facility alone due to distance. | 123.4233 | 324.456 | 0.362 | 0.84 |
| I have to take public transportation to visit healthcare. Facility, but there is often a lack of transportation. | 123.2869 | 332.427 | 0.133 | 0.846 |
| I do not have post-delivery follow-up support. | 124.0057 | 319.345 | 0.351 | 0.84 |
| High-quality health services are not present in public hospitals. | 124.1193 | 320.613 | 0.314 | 0.841 |
| The pressure to have a son makes my pregnancies difficult. | 123.3835 | 323.565 | 0.376 | 0.839 |
| I was unable to read the brochures given by doctors about maternal healthcare during pregnancy. | 123.2131 | 320.704 | 0.45 | 0.838 |
| I do not know different methods of contraception. | 123.2699 | 326.044 | 0.303 | 0.841 |
| I was unaware of the significance of multivitamins during pregnancy. | 123.2131 | 320.362 | 0.429 | 0.838 |
| Men in the family are not concerned about women's health during pregnancy. | 123.8381 | 341.515 | -0.065 | 0.852 |
|  |  |  | **Cronbach's alpha 0.844** |  |

### Regression Analysis

Table 3 represents the model summary and ANOVA results. The model summary shows the strength of the relationship between the model and the dependent variable, namely the model’s fitness, which is 44.90%. In this connection, several factors leading to maternal and perinatal health vulnerabilities include poverty and gender inequality. However, there is a significant variance between MHV and gender inequality and poverty. 05 p-value is a cutoff value that shows the possibility of acceptance of the null hypothesis. MHV and gender inequality and poverty are two separate things, but they are interdependent in the context of women.

**Table 3.** The Multiple Regression Analysis (Model summary and ANOVA)

| Model Summary |  |  |  |  |  |
| --- | --- | --- | --- | --- | --- |
| R |  | R Square | Adjusted R Square | Std. Error of the Estimate |  |
| .672<sup>a</sup> |  | 0.452 | 0.449 | 0.74240223 |  |
| ANOVA |  |  |  |  |  |
|  | Sum of Squares | df | Mean Square | F | Sig. |
| Regression | 158.645 | 2 | 79.322 | 143.919 | .000<sup>b</sup> |
| Residual | 192.355 | 349 | 0.551 |  |  |
| Total | 351 | 351 |  |  |  |

a. Predictors: (Constant), Gender Inequality, Poverty.
b. Dependent Variable: Maternal Health Vulnerabilities.

Table 4 represents the results of regression Coefficients, showing the significant level of poverty and gender inequality in the prediction of MHV. The Beta value shows the contribution of each independent variable separately. In this study, poverty predicts MHV positive and significant (65.60% and .000). In contrast, gender inequality predicts MHV positive and significant (10.50% and .000). Thus, in MHV, gender inequality and poverty are two different terms that tend to complement each other by acting as risk or protective factors, respectively, both at the individual level and at the health system—further sig. Values also explain the status of hypotheses H1 and H2, which are supported in the study.

**Table 4.** Unstandardised and standardised Coefficients.

| Coefficients |
| --- |

|  | **Unstandardised Coefficients** | **Std. Error** | **Standardised Coefficients** | **t** | **Sig.** |
| --- | --- | --- | --- | --- | --- |
| (Constant) | -5.92E-17 | 0.04 |  | 0 | 1 |
| Poverty | 0.656 | 0.04 | 0.656 | 16.517 | 0 |
| Gender Inequality | 0.105 | 0.04 | 0.105 | 2.637 | 0.009 |

## Discussion and conclusion

Reducing maternal mortality has become a global health priority as it affects women disproportionately in low and middle-income countries. Many of these deaths are preventable [23]. Maternal health has become a challenge for Pakistan, which faces a maternal health crisis with a high MMR. The prevalent societal beliefs and perceptions, household power structures and misinformation significantly impact maternal health [24]. The results of this study reveal that different levels within the SEM influence factors affecting maternal health vulnerabilities. These factors include sociodemographic characteristics of the women and social and community aspects.

Women’s vulnerabilities in rural areas begin from the micro and mesosystem: poverty, gender inequality, and cultural practices disempower women and pose numerous barriers to enhancing their social status. The women lack education and awareness about reproductive health. This is even though education and awareness about reproductive and maternal health can play a central role in smoothing ANC, which is central to promoting a positive pregnancy experience and improving maternal and child survival [25]. The critical steps of ANC include risk diagnosis, pregnancy management and prevention of pregnancy-related diseases, health education and health promotion [26]. The participants in this study showed a lack of health awareness and information about ANC. Most participants, i.e., 49%, were not literate, while 19% had primary-level education. Only 6% of participants had matric, and 4% had intermediate-level education.

Thus, most of the women were unable to read and comprehend information about ANC, the use of contraceptives, the importance of follow-ups, maintaining diet plans, reading and understanding brochures, knowledge about modern healthcare facilities, and the extent of contraceptives.

Patriarchy, supported by socio-cultural norms, is another element within the micro, meso and exosystem impacting MHV. In rural areas, the uptake of ANC is widely influenced by social and cultural norms. Inadequate and poor-quality services and social and cultural orientations limit women’s access to quality maternal health services [4]. Results of this study also reflect that women’s regular visits to healthcare facilities and follow-ups were greatly influenced by the distance of the health facility from their residence; a more significant distance complicated the situation further. Since men make substantial decisions, women’s access to ANC services was also subject to men’s approval.

Women’s low status, poverty, limited literacy, and inaccessible, inadequate and costly healthcare services influence MHV [27]. Results of this study show that poverty is the main factor influencing ANC practices in the villages; the women in this study were less likely to maintain the healthy diet required during pregnancy due to financial constraints such as milk consumption, proper food, or vitamin intake. Women also preferred home delivery because their families could not afford hospital expenses. Poverty is a significant factor in the micro, meso, and exosystems of SEM that influence MHV.

The growing population and poor maternal health outcomes in Pakistan require immediate response and improved strategies. For example, gains cannot be made by investing in the health sector alone; social indicators of women also need close attention. Fundamental issues about women’s status, empowerment, education, hygiene and sanitation must also be considered [28, 29].

There is a lack of gender-sensitive policies and initiatives on the macro level to address MHV in rural areas and remote communities where women are in disadvantageous positions. Maternal health outcomes can be improved with robust interventions targeted to reduce gender inequality in accessing healthcare. The World Health Organization has set a goal to decrease the global MMR to 70 maternal deaths per 100,000 live births by 2030 [30]. For this, WHO is working to record maternal mortality through increased research evidence, setting global standards, providing evidence-based clinical and pragmatic guidance, and giving technical support to member countries in developing and implementing effective policies and programs [4].

Moreover, pregnancy-related awareness programs, the provision of skilled birth attendants and access to antenatal care contribute to safer pregnancies and childbirth. To address the needs and priorities of women and girls, inequalities must be removed in accessing quality reproductive, maternal, and healthcare services for newborn children, along with the factors causing maternal mortalities, morbidities, and related disabilities.

The SEM helped to enhance an understanding of how different factors at personal, community and societal levels influence individuals’ health-seeking behaviour and increase vulnerabilities. This study concludes that gender inequality and poverty are well entrenched in rural areas, resulting in poor ANC practices. We suggest addressing the needs and priorities of the women residing in less privileged areas. This can only be possible if socio-economic and geographical factors are considered and women are given improved access to financial and physical resources. This study is limited to some villages in northern Sindh; more research on MHV is required in other less privileged areas of Pakistan where patriarchy is well entrenched and women’s access to public spheres is restricted.

## Disclaimer

None

## Data Availability

All relevant data are within the manuscript and its Supporting Information files.

## Acknowledgement

We would like to thank our funders, the Academy of Medical Sciences UK, for their support of the study and Dr Zahid Memon for helping us obtain ethics approval.

